# METABOLIC DYSFUNCTION-ASSOCIATED STEATOTIC LIVER DISEASE IN PATIENTS WITH INFLAMMATORY BOWEL DISEASE: ASSOCIATION WITH BODY MASS INDEX AND DISEASE SEVERITY

**DOI:** 10.64898/2026.10.12.26365296

**Authors:** A.A. Antoniv, N.A. Antoniv

**Author notes:** Corresponding author: Antoniv Alyona Andriivna, Bukovinian State Medical University, 2 Teatralna Square, Chernivtsi 58002, Ukraine. **Connection of the publication with planned research work** The study was undertaken within the departmental research project “Features of the Combined Course of Internal Organ Diseases: Mechanisms of Mutual Aggravation and Approaches to Pharmacological Correction” (state registration number 0124U002435).

## Abstract

Whether body mass index (BMI) can meaningfully distinguish inflammatory bowel disease (IBD) patients with concomitant metabolic dysfunction-associated steatotic liver disease (MASLD) was examined in a retrospective hospital series. The analysis covered 400 records from 2023–2024 with verified ulcerative colitis or Crohn’s disease and sufficient information on anthropometry, liver status, and intestinal disease activity. Normal BMI was documented in 118 patients, 176 were overweight, and 106 had obesity. MASLD classification required documented hepatic steatosis in the appropriate cardiometabolic setting; ultrasonography provided the principal evidence of steatosis, while laboratory findings, non-invasive indices, and elastography were considered when available. Ulcerative colitis activity was based on the Mayo Score and Crohn’s disease activity on CDAI. Overall, 181 patients (45.3%) met MASLD criteria. The corresponding frequencies across the three BMI strata were 20.3%, 49.4%, and 66.0% (χ^2^=49.29; p<0.001). Severe IBD was recorded in 6.8%, 14.8%, and 32.1%, respectively (χ^2^=39.45; p<0.001). Relative to normal BMI, the unadjusted odds of MASLD were 3.83-fold higher with overweight (95% CI 2.24-6.55) and 7.62-fold higher with obesity (95% CI 4.17-13.91). Thus, BMI categories identified groups with distinctly different burdens of both MASLD and severe IBD. Because treatment exposure, disease duration, and other covariates were not included in an adjusted model, the observed differences should be interpreted as associations rather than causal effects.

## Introduction

Abnormal liver findings in inflammatory bowel disease (IBD) are interpreted against a changing clinical background. In ulcerative colitis and Crohn’s disease, weight and body composition may vary with inflammatory activity, nutritional intake, treatment exposure, and physical activity. Hepatic steatosis in this setting therefore cannot be viewed only as an incidental imaging finding or as a conventional metabolic disorder detached from the intestinal disease [1,2].

The metabolic dysfunction-associated steatotic liver disease (MASLD) framework is useful for this clinical overlap because it defines disease positively: steatosis must coexist with cardiometabolic risk. Current European recommendations extend the evaluation beyond the presence of liver fat and call for assessment of accompanying metabolic abnormalities and stepwise estimation of fibrosis risk [3,4].

Published data confirm that fatty liver is frequent in IBD, but they do not indicate one uniform prevalence. Zamani et al. synthesized 44 studies and reported 30.7%, whereas a global meta-analysis released in 2026 found 25.4% among more than 1.5 million individuals. Differences of this magnitude are plausible because hospital versus outpatient recruitment, metabolic characteristics of the sample, and the method used to detect steatosis can all shift the observed frequency [5,6].

Excess adiposity explains only part of the overlap. MASLD has also been documented in lean individuals with IBD, and one analysis retained an independent association with IBD after adjustment for conventional metabolic variables. This supports a broader model in which metabolic susceptibility may coexist with gut-related processes, including barrier impairment, dysbiosis, altered bile-acid signaling, and persistent inflammatory activation. Recent reviews of the gut-liver axis in IBD likewise emphasize the interaction of intestinal barrier dysfunction, microbial signaling, and metabolic abnormalities rather than a single dominant pathway [2,7,8].

The clinically relevant question is therefore not simply whether obesity and MASLD coexist. It is whether routine BMI categories identify patients in whom liver involvement and a more severe intestinal course cluster within the same clinical setting. We examined this question in a single hospital cohort for which both hepatic and IBD-activity data were available.

### The aim of the study

The study compared MASLD occurrence and IBD severity across predefined BMI strata and calculated unadjusted pairwise odds of MASLD for those strata.

### Object and research methods

The study dataset comprised 400 inpatient charts from the gastroenterology and internal medicine departments of Chernivtsi Regional Clinical Hospital and Chernivtsi Emergency Hospital. Admissions from 2023–2024 were eligible when ulcerative colitis or Crohn’s disease had been verified and the chart contained height, weight, liver-assessment data, and sufficient information to classify intestinal disease activity. Age ranged from 24 to 67 years (mean 45.8 ± 10.4 years). There were 216 men (54.0%) and 184 women (46.0%).

Height and weight recorded in the charts were used to calculate BMI. Participants were assigned to one of three categories: 18.5–24.9 kg/m^2^ (normal BMI; n=118), 25.0–29.9 kg/m^2^ (overweight; n=176), or ≥30.0 kg/m^2^ (obesity; n=106). The groups were similar in age, sex distribution, and the relative proportions of ulcerative colitis and Crohn’s disease (all p>0.05).

IBD verification required concordant clinical, laboratory, endoscopic, and histological documentation. Ulcerative colitis activity was obtained from the Mayo Score, whereas Crohn’s disease activity was taken from the Crohn’s Disease Activity Index (CDAI). For analyses pooling the two diseases, the original disease-specific activity estimates were aligned into mild, moderate, and severe categories without replacing the underlying scores [1].

MASLD status was reconstructed from the information available in each chart. A patient was assigned to the MASLD group when hepatic steatosis had been documented together with cardiometabolic criteria compatible with the current definition. Abdominal ultrasonography was the primary source of steatosis evidence. Anthropometric and biochemical data were reviewed alongside imaging; Hepatic Steatosis Index, Liver Fat Score, non-invasive fibrosis indices, and liver elastography were used as supplementary information when available. The routine hepatic panel included alanine aminotransferase, aspartate aminotransferase, gamma-glutamyl transferase, alkaline phosphatase, bilirubin fractions, and albumin [3,4].

Charts were omitted when another major condition could substantially distort the interpretation of liver findings or systemic inflammation. These conditions comprised viral hepatitis B or C, alcohol-related liver disease, autoimmune liver disease, malignancy, severe renal failure, decompensated cardiovascular disease, and infectious colitis.

Each patient contributed one observation to the analysis. Categorical data are presented as n (%), and age as mean ± standard deviation. Pearson’s χ^2^ test was used for the BMI-by-MASLD and BMI-by-IBD-severity contingency tables. Crude odds ratios (ORs) with 95% confidence intervals (CIs) were obtained from pairwise 2×2 comparisons. A two-sided p value <0.05 defined statistical significance. Calculations were performed in Statistica for Windows 10.0 and Microsoft Excel 2016; the main contingency-table results were also recalculated directly from the reported cell counts.

Ethical oversight was provided by the Biomedical Ethics Committee of Bukovinian State Medical University (protocol No. 10, June 17, 2026). The source records documented informed consent for clinical investigations and research use of clinical information, and the study followed the principles of the Declaration of Helsinki and Good Clinical Practice.

### Research results and their discussion

Of the 400 records, 282 (70.5%) belonged to patients whose BMI was at least 25 kg/m^2^, and 181 patients (45.3%) met MASLD criteria. Table 1 shows a pronounced ordering by BMI: MASLD was least frequent in the normal-BMI stratum and most frequent in the obesity stratum (χ^2^=49.29, df=2; p<0.001).

**Table 1.**
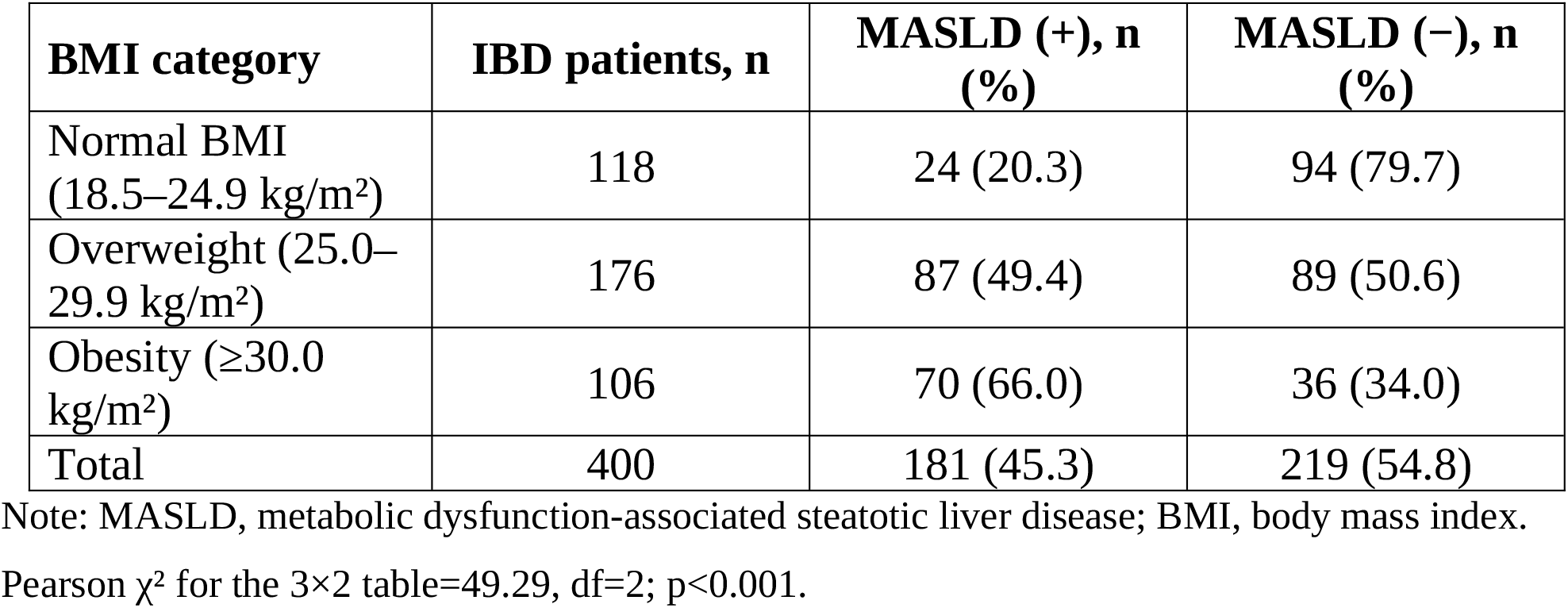
Distribution of MASLD within BMI strata.

The largest absolute change occurred before the obesity threshold. Moving from normal BMI to overweight corresponded to an increase in MASLD frequency from 20.3% to 49.4%, or 29.1 percentage points. The subsequent increase from overweight to obesity was smaller, at 16.6 percentage points. Thus, in this cohort, a substantial rise in liver involvement was already apparent in the overweight range.

A similar ordering was seen for intestinal disease severity (table 2). Mild IBD became less common across increasing BMI categories, whereas the proportion classified as severe rose from 6.8% in the normal-BMI group to 14.8% in the overweight group and 32.1% in the obesity group. The overall distribution differed significantly among the three BMI strata (χ^2^=39.45, df=4; p<0.001).

**Table 2.** IBD severity profile by BMI stratum.

| **IBD severity** | **Normal BMI (n=118)** | **Overweight (n=176)** | **Obesity (n=106)** |
| --- | --- | --- | --- |
| Mild | 67 (56.8%) | 71 (40.3%) | 24 (22.6%) |
| Moderate | 43 (36.4%) | 79 (44.9%) | 48 (45.3%) |
| Severe | 8 (6.8%) | 26 (14.8%) | 34 (32.1%) |

Note: ulcerative colitis activity was classified from the Mayo Score and Crohn's disease activity from CDAI; both were subsequently expressed as mild, moderate, or severe for pooled analysis.
Pearson $\chi^2$ for the 3×3 table=39.45, $df=4$ ; $p<0.001$ .

The pairwise estimates in table 3 quantified the same pattern. Compared with normal BMI, overweight corresponded to an OR of 3.83 for MASLD and obesity to an OR of 7.62; obesity also differed from overweight (OR=1.99). These are crude estimates. They describe how MASLD and BMI categories co-occurred in the dataset and should not be interpreted as independent effects of body weight.

**Table 3.** Pairwise crude odds of MASLD across BMI categories.

| **Comparison** | **OR** | **95% CI** | **p** |
| --- | --- | --- | --- |
| Overweight vs normal BMI | 3.83 | 2.24–6.55 | <0.001 |
| Obesity vs overweight | 1.99 | 1.21–3.28 | 0.0066 |
| Obesity vs normal BMI | 7.62 | 4.17–13.91 | <0.001 |

Note: OR, odds ratio; CI, confidence interval. Values were calculated from the cell counts in table 1 without covariate adjustment.

The observed MASLD frequency of 45.3% is above the pooled values reported in large IBD evidence syntheses, including 30.7% and 25.4% [5,6]. This difference does not necessarily indicate inconsistency. The present sample was entirely hospital-based, and 70.5% of patients were above the normal BMI range. Both characteristics make a higher burden of metabolic comorbidity plausible. The 45.3% figure should therefore be regarded as a characteristic of this selected cohort rather than a prevalence estimate for the regional IBD population.

The normal-BMI stratum is equally informative. MASLD was found in 24 of 118 patients despite body weight being within the normal range. Reports from lean IBD populations show a similar principle: steatotic liver disease can occur without overweight and may remain linked to IBD after conventional metabolic factors are taken into account. BMI therefore ranks patients by observed frequency, but it cannot serve as a rule for excluding MASLD [7].

IBD severity changed in parallel with the BMI gradient. The share of severe disease increased across the same categories in which MASLD became more frequent. This pattern is clinically relevant, but the design does not establish direction. Adiposity may accompany a less favorable inflammatory-metabolic environment, while active IBD, corticosteroid exposure, lower physical activity, dietary changes, and altered body composition may themselves influence weight. The finding is therefore best interpreted as a concurrent pattern within the cohort rather than evidence that higher BMI causes more aggressive intestinal disease.

Several mechanisms could provide biological context for the coexistence of IBD and MASLD. The literature points to impaired intestinal barrier function, microbiome-derived signals, disturbed bile-acid metabolism, inflammatory mediators, insulin resistance, and visceral adiposity. None of these pathways was measured directly in this dataset, so they cannot be claimed as mediators of the associations reported here [2,8].

For clinical practice, BMI is most useful as an indicator to intensify review rather than as a diagnostic threshold. Overweight and obesity identified groups in which MASLD was common, supporting careful assessment of liver biochemistry, cardiometabolic risk, and available measures of steatosis or fibrosis. Prospective IBD data link MASLD and its cardiometabolic components with subsequent cardiovascular events. In addition, current MASLD literature emphasizes fibrosis assessment as a central component of long-term risk stratification. At the same time, the 20.3% frequency in the normal-BMI stratum shows why hepatic evaluation should still be guided by biochemical, metabolic, and imaging clues regardless of body size [9,10].

Interpretation is limited by the retrospective design, which leaves residual confounding and provides no temporal sequence between BMI, MASLD, and intestinal activity. Hospital recruitment may also select for more active or complicated IBD. Treatment exposure, disease duration, waist circumference, diabetes, dyslipidemia, and other potential covariates were not recorded consistently enough for multivariable adjustment. Liver assessment followed routine clinical practice rather than a uniform research protocol. Finally, the reported ORs are unadjusted and require confirmation in prospectively assembled cohorts with standardized liver phenotyping.

## Conclusions

1. MASLD frequency differed markedly across BMI strata, with the largest absolute increase occurring between normal BMI and overweight rather than only after the obesity threshold was reached.
2. The distribution of IBD severity changed in the same direction: severe disease became progressively more common across higher BMI categories, although the retrospective design does not establish temporal order or causality.
3. BMI may help prioritize metabolic and hepatic assessment in IBD, but it should not be used to exclude MASLD because clinically relevant liver involvement was also present in patients with normal BMI.

### Prospects for further research

Future studies should use prospective, protocol-based recruitment to determine whether BMI adds predictive information beyond established metabolic and IBD-related variables. Such work should include waist-based measures of adiposity, treatment history, disease duration, standardized quantification of steatosis and fibrosis, and validated measures of IBD activity. Adding inflammatory biomarkers, intestinal permeability indices, and microbiome characteristics would allow multivariable evaluation of metabolic and intestine-derived pathways.

## Data Availability

The de-identified data supporting the findings of this study are available from the corresponding author upon reasonable request, subject to applicable ethical and data protection requirements.

## Ethics approval and consent to participate

The study was approved by the Biomedical Ethics Committee of Bukovinian State Medical University (protocol No. 10, June 17, 2026). The source medical records documented informed consent for clinical investigations and research use of clinical information. The study was conducted in accordance with the principles of the Declaration of Helsinki and Good Clinical Practice.

## Data availability

The data supporting the findings of this study are available from the corresponding author upon reasonable request. The data are not publicly available because they contain information derived from medical records and are subject to institutional and ethical restrictions.

## Author contributions

A.A. Antoniv: work concept and design; data collection and analysis; responsibility for statistical analysis; writing the article; critical review; final approval. N.A. Antoniv: data collection and analysis; writing the article; critical review; final approval. Both authors reviewed and approved the final manuscript.

